# A Poorly Understood Disease? The Unequal Distribution of Excess Mortality Due to COVID-19 Across French Municipalities^*^

**DOI:** 10.1101/2020.07.09.20149955

**Authors:** Paul Brandily, Clément Brébion, Simon Briole, Laura Khoury

**Author notes:** We would like to thank Philippe Askénazy, Luc Behaghel, François Fontaine, Clément Malgouyres, Éric Maurin, Dominique Meurs as well as participants of the“Chaire Travail”seminar at the Paris School of Economics for their valuable comments. We thank as well the *CASD* for providing us access to the data. E-mail addresses.

## Abstract

While COVID-19 was already responsible for more than 500,000 deaths worldwide as of July 3, 2020, very little is known on the socio-economic heterogeneity of its impact on mortality. In this paper, we combine several administrative data sources to estimate the relationship between mortality due to COVID-19 and poverty at a very local level (i.e. the municipality level) in France, one of the most severely hit country in the world. We find strong evidence of an income gradient in the impact of the pandemic on mortality: it is twice as large in the poorest municipalities compared to other municipalities. We then show that both poor housing conditions and higher occupational exposure are likely mechanisms. Overall, these mechanisms accounts for up to 60% of the difference observed between rich and poor municipalities.

## 1 Introduction

While COVID-19 is directly responsible for at least 521,000 deaths around the globe, as of July 3,^1^ we know very little on the heterogeneity of its impact on mortality across regions and individuals. In particular, while an emerging literature in the US shows that ethnic minorities exhibit higher levels of COVID-19 prevalence (Almagro and Orane-Hutchinson, 2020; Borjas, 2020; McLaren, 2020), the evidence on the heterogeneous impact of COVID-19 on mortality across income groups remains scarce.

In this paper, we explore this issue in the context of France, one of the most severely hit country in the world. We combine several administrative data sources to estimate the relationship between excess mortality due to COVID-19 and poverty at the municipality level, a very detailed geographic scale (1,600 inhabitants and 15 sq.km per municipality, on average). To the best of our knowledge, this paper is the first to rely on exhaustive datasets at such level of precision in order to study this socio-economic aspect of COVID-19. Accurately estimating this relationship is one of the main challenges faced by the literature because of the limited availability and reliability of public data on mortality directly attributable to COVID-19.^2^ We circumvent this issue by using exhaustive death records provided by the French National Statistical Institute (INSEE) on a daily basis and we focus on excess mortality, broadly defined as the deviation in mortality with respect to a reference (pre-COVID) period.

We also take advantage of a quasi experimental setting to identify the causal impact of COVID-19 on mortality at the municipality level. The evolution of mortality across time can be affected by confounding factors not directly related to COVID-19. One obvious factor relates to the very special life conditions implied by the lockdown, but health-related factors due to year-to-year variations in seasonal diseases may also have an impact on excess mortality.^3^ To overcome this issue, we exploit the fact that the lockdown was uniformly implemented over the French territory on March 17 (up until May 11) whereas the spread of the epidemic was then very heterogeneous across regions. In particular, we can distinguish high-infection areas, where COVID-19 was already widely spread when the lockdown started, from low-infection areas, where the prevalence of COVID-19 was very low but where life conditions were completely identical to those of high-infection regions.^4^ We build on this quasi experimental setting by employing a triple-difference strategy which consists in comparing the evolution of mortality between rich and poor municipalities located in high-infection regions vs. in low-infection regions. Our analysis compares municipalities within urban areas to investigate the role of very local socio-economic determinants of the spread of the virus. We see this identification of the causal impact of poverty on COVID-19 related mortality as an improvement on the current literature.

We find strong evidence of a positive relationship between municipalities’ mortality due to COVID-19 and their poverty level. In high-infection regions, the pandemic caused a 50pp increase in mortality in most municipalities (relative to low-infection areas), but this increase actually reaches 88pp for municipalities of the poorest quartile (i.e. municipalities where the median household income is below the 25^*th*^ percentile of the national income distribution). In these regions, the impact of the pandemic is therefore twice as large in poor municipalities. By contrast, the comparison of rich and poor municipalities in low-infection regions reveals no significant difference in the evolution of mortality. These results are robust to alternative definitions of poverty and mortality which focus on the elderly or use the death toll rather than the death ratio.

Based on the same methodological approach, we then explore potential mechanisms explaining the causal relationship between mortality and poverty observed at the municipality level. An extensive literature provides evidence of substantial health inequalities across individuals of different socio-economic status (Adler and Rehkopf, 2008; Glied et al., 2012). A recent work by Wiemers et al. (2020) confirms a higher incidence of severe complications from COVID-19 among poor individuals. While we acknowledge the key role that health factors play at the individual level, we focus on two complementary mechanisms related to disease transmission channels at the municipality level.^5^ Combining social security records (DADS) that provide detailed information on occupation at the individual level with a recent survey on working conditions, we first build two complementary measures of occupational exposure at the municipality level, based (i) on the frequency of social contacts under regular working conditions and (ii) on the share of jobs in sectors authorized to continue their activity during the lockdown. We then match this dataset with additional data provided by the French national institute of statistics on the share of overcrowded housing units in every French municipalities. Finally, to investigate the potential interaction between occupational exposure and housing conditions, we exploit census data to compute the share of multi-generational households that include at least one worker and an elderly person (over 65 y.o.) in municipalities with more than 2,000 inhabitants.

A preliminary analysis confirms that all our indexes measuring occupational exposure and poor housing conditions strongly relate to municipalities’ poverty level. Further analysis shows that the share of overcrowded housing unit unambiguously increases mortality due to the pandemic. The evidence is more mixed regarding the effects of occupational exposure: while the frequency of social contacts acts as a channel of disease transmission, the relationship between the share of authorized jobs during the lockdown and excess mortality is not significant. Finally, the share of multi-generational households significantly and positively relates to excess mortality, suggesting that the effects of poor housing conditions and occupational exposure on mortality may partially operate through working-age individuals transmitting the virus to more vulnerable individuals in their household. A Oaxaca-blinder decomposition finally reveals that, taken together, these mechanisms account for up to 60% of the differential mortality pattern observed between rich and poor municipalities located in high-infection regions.

We make several contributions to the emerging literature on socio-economic aspects of COVID-19 consequences. A first strand of this literature highlighted the unequal consequences of the COVID crisis on individuals’ labor market outcomes (Adams-Prassl et al., 2020; Alstadsæter et al., 2020; Borjas and Cassidy, 2020; Fairlie et al., 2020; Montenovo et al., 2020; Yasenov, 2020). These papers tend to show that the crisis primarily affected vulnerable groups in terms of income or job loss, especially ethnic minorities. A more closely related literature examines the unequal distribution of COVID-19 infection and related mortality across ethnic groups (Borjas, 2020; McLaren, 2020; Sa, 2020). Our main contribution is to provide clear evidence of a substantial income gradient in the impact of the pandemic on mortality. As compared to previous papers, we are able to base our analysis on exhaustive data available at a very local level for a very large population. We also develop a triple difference estimation strategy which allows us to isolate the effect of COVID-19 from that of lockdown on excess mortality. Finally, we highlight the key role played by poor housing conditions and occupational exposure in the unequal impact of COVID-19 on mortality.

We also contribute to the growing literature studying the factors influencing the spread of COVID-19. This literature studies the role of occupational exposure (Almagro and Orane-Hutchinson, 2020; Lewandowski, 2020), economic activities’ concentration (Ascani et al. (2020)) or mass protests (Dave et al. (2020)) in the spread of the epidemic. In our paper, we build an index of social contacts at the occupational level and we provide causal evidence on the effect of occupational exposure on mortality due to COVID-19. We also highlight the critical role played by housing conditions in the spread of the epidemic and its impact on mortality. While most previous papers exploit survey data, we are able to build on administrative data available at a very local level for the whole population of mainland France. Finally, to the best of our knowledge, our paper is also one the first to study potential transmission channel using mortality rather than COVID-19 detected cases.

The remainder of the paper is organized as follows: section 2 describes our main data sets and the construction of our variables of interest. In section 3, we present our empirical strategy before estimating the link between poverty and excess mortality due to COVID-19 in section 4. Given these results, in section 5 we explore potential mechanisms. We also provide robustness checks for our main results in section 6 and section 7 concludes.

## 2 Data and Measurement

In this paper, we gather various data sets using municipality-level identifiers. In France, municipalities are small administrative units of 1,600 inhabitants and 15.3 sq.km on average (as of 2014); there are about 35,000 of them in 2020. Our analysis compares municipalities *within* urban areas. Urban areas are groups of neighboring municipalities (see Appendix A for more details) defined by the French National Statistical Institute (INSEE). There are 706 urban areas in France with a median number of municipalities of 89 and a median population size of 2,358 inhabitants. The majority of our sources are exhaustive administrative data sets made available by INSEE. Below, we briefly mention the main data sources we use and we also provide more details in Appendix A.

### 2.1 Mortality

To measure mortality to COVID-19 at the municipality level, we rely on daily counts of deaths. For each day from January to the end of May, INSEE reports death records including municipality and place (hospital or clinic, home, care home, etc.) of death as well as a set of individual-level characteristics such as sex, date of birth and municipality of residency. These files do not record the cause of death and, in particular, do not identify COVID-related death. This is one reason why most of our analysis is based on excess mortality (cf. next paragraph below) comparing April 2020 records to their 2019 and 2018 counterparts. In the emergency of the COVID-crisis, INSEE made the data set available at a high frequency rate: this has induced potential measurement errors although INSEE applies corrections at each iteration. We recognize this limitation and update our data as often as possible. Note that, in theory, municipalities have about one week to inform INSEE about new death. Most of our analysis focuses on the excess mortality in the month of April - because it captures the peak of the epidemic in France. As the version of the data that we are currently using got downloaded on June 30, our measures should be accurate.^6^

We do not use information relative to COVID-19 testing both because such data is not available at the municipality level (to our knowledge) but also because testing is unlikely to be randomly allocated, especially in the case of France that ranks among the worst OECD countries in number of tests per inhabitant (Scarpetta et al. (2020); cf. also Foucart and Horel (2020) on the French context and Borjas (2020) on tests allocation in New York City).

#### Excess mortality

Our main outcome variable is the excess mortality in April 2020. It corresponds to the ratio between death rate in April 2020 and the average of the death rate in April 2018 and 2019.^7^ Because we use the same population value (recorded in 2014) for the computation of all three death rates, it translates into the ratio of the number of deaths in April 2020 and the average of the number of deaths in April 2019 and 2018, as follows:

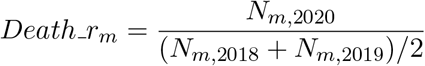

where *N*_*m,y*_ is the number of deaths in municipality *m* in April of year *y*.

This definition of the excess mortality rate follows the guidelines defined by the Centre for Epidemiology of Medical Causes of Death of the French National Institute of Health and Medical Research (INSERM, CepiDc) available here. As it is a relative measure, it takes into account population size.^8^

The French National Institute of Health and Medical Research has used alternative measures in previous studies (SPF, 2019), such as the difference in the number of deaths:

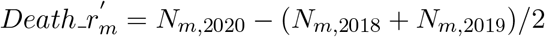

We therefore provide a robustness check using this definition, and results hold (Table A2 of Appendix 3).

It has been shown that likelihood to die from COVID-19 is much higher among people over 65 years old (Wu and McGoogan, 2020).^9^ However, to be as agnostic as possible, we use the total count of deaths, without focusing on the elderly. We also provide, in Section 6, the coefficient of our main regression for different age groups (Figure A2 of Appendix 3).

Finally, we favor the use of all-cause excess mortality data instead of data on confirmed cases of COVID-19 to measure COVID-19 related mortality, because the latter suffers from several biases: (i) as previously mentioned, testing was only scarce in France, and not randomly implemented; (ii) people dying from COVID-19 may not be accurately identified (if dying at home for example). According to Silverman et al. (2020) projections, almost 80% of cases of COVID-19 in the US were never diagnosed. One last drawback of the data on confirmed cases of COVID-19 in France is that it is only available for people who died at the hospital, and at the *département* level. We believe that conducting the analysis at the municipal level is crucial, as it allows a more careful investigation of the socio-economic determinants of the mortality due to COVID-19. Indeed, if the location of a cluster within a country may be due to non-relevant reasons (e.g. the organisation of a large event), the way it spreads within a limited area may more likely be influenced by socio-economic factors. Note that using all-cause excess mortality could also create potential biases as it captures both the effect of COVID-19 and of the lockdown, and because it can be sensitive to random variations (Le Bras, 2020). We address these issues in two ways: (i) by measuring excess mortality over a month, not to be too sensitive to day-to-day random variations in the number of deaths; (ii) by using an empirical strategy that isolates the effect of COVID-19 from that of the lockdown (see Section 3).

#### High- and low-infection regions

The French government decided to implement a national lockdown policy from March 17 to May 11. During this period, lockdown conditions were strictly identical across the territory. However, the prevalence of COVID-19 was already much higher in some regions when the lockdown started. As a consequence, more restrictive measures were imposed in high-infection *départements* when the government decided to implement a gradual lifting of COVID-19-related restrictions after May 11.^10^ In practice, the Ministry of Public Health provided a map differentiating red (high-infection) and green (low-infection) *départements* on May 7 (cf. Figure A1 in the Appendix). This map distinguishes high- and low-infection areas on the basis of: (i) the share of ER visits for a COVID-19 suspicion; (ii) the occupancy rate of intensive care beds relative to the initial one; (iii) the testing rate relative to the needs. High-infection *départements* roughly correspond to 30% of mainland French *départements* and are located in the North-Eastern quarter of the country. As further explained in Section 3, our analyses will compare municipalities within high and low-infection areas based on the definition by the Ministry of public health, allowing us to isolate the effect of COVID-19 from that of the lockdown.

### 2.2 Poverty

Our main independent variable is a measure of poverty at the municipal level. We rely on information on the standard of living from the *Filosofi* database in 2014. This database, built from tax data, contains information on poverty and various sources of income at a local level (see Appendix A for more details). The concept of *standard of living* corresponds to the household disposable income divided by consumption units (using the OECD scale), to account for the size of the household. Based on this income information, we define two poverty dummies that respectively take the value one if the median standard of living of the municipality is: (i) below the median of all municipalities (weighted by population size); (ii) below the first quartile of the weighted national distribution.

### 2.3 Labor Market Conditions

One of the main novelties of this paper is to use administrative data to test whether labor market conditions explain the differential in excess mortality between poor and rich municipalities. We focus on two main characteristics of jobs that we expect to be key vectors for spreading the disease: (i) whether the worker kept on going to her workplace during the lockdown; (ii) whether the job involves being in contact with others.

To measure the first dimension, we use a decree issued by the French government on March, 23rd 2020 (source). The decree gives an explicit list of sectors that could remain open during the lockdown. We use the sectors’ names listed in the decree to match them with their corresponding official classification (at 4 digits). Such process is noisy by nature, though to a limited extent. We then recover individual information about sector of employment using either the DADS or the Census, depending on our analyses. For each municipality we compute the share of workers that are employed in firms entitled to remain open.^11^ This measure is a proxy of our feature of interest as some of these workers did not keep on going to their workplace during the lockdown (some may have worked from home, some of their firms may have temporarily closed their doors, etc.). In other words, such measure captures an “intention to treat” and is, to our knowledge, the best available approximation.

As for the second dimension, our proxy is based on the question “In your job, are you in direct contact with the public” that is available in a survey called DEFIS (see Appendix A). For each occupation code (at the 3-digit level), we compute the share of workers answering “Often” (v.s. “Sometimes” or “Never”). Using the DADS, we then compute the average of this index at the municipality level based on the occupation distribution of workers living in each municipality.

### 2.4 Housing Conditions

Our last dimension of interest is the relation between housing conditions and excess mortality due to COVID-19. We use the average share of housing units that are overcrowded provided by INSEE at the municipality level. Overcrowded accommodations are those that have less than “one living room, one room for each couple, one room for each other adult aged 19 or older, one room for two children if they are of the same sex or are under 7 years of age, and one room per child otherwise”.

### 2.5 Interaction between Labor Market and Housing Conditions

To investigate the interactions between both mechanisms, we finally build an index at the municipality level, based on the partial census data files available for municipalities with more than 2,000 inhabitants. We compute the share of households with both an elderly person (over 65 y.o.) and a worker that is younger by at least 18 years (“multi-generational households” hereafter). This variable is meant to capture the fact that having different generations living in the same apartment increases the likelihood to get infected, and to potentially die for the elderly. This likelihood increases further if at least one of the younger members of the household works, as it potentially multiplies her social interactions.

## 3 Empirical strategy

Our main method of identification is described in Equation 1. This model is a triple-difference regression that estimates the change in mortality rate at the municipality level over three dimensions: (i) time; (ii) the infection intensity of COVID-19 and (iii) income. The time dimension consists in comparing the number of deaths in April 2020 with the number of deaths in April 2018 and 2019. The second dimension accounts for the contamination intensity of COVID-19 across municipalities. It is measured by the high and low-infection *départements* defined by the government when it started lifting COVID-19-related restrictions (see Section 2 on data description). We use these two groups to classify cities on the second dimension and we define the variable *Infect*_*d*_ that takes the value one for municipalities in high-infection *départments*. The third and last dimension divides municipalities according to their median standard of living. We discuss the cut-off that we use to split the national distribution of this measure into rich and poor municipalities in the following section. In Equation 1, the variable *Poor*_*m,ua,d*_ is a dummy that takes the value one if municipality *m* of urban area *ua* and *département d* is below this cut-off.

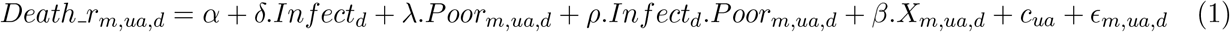

where *Death_r*_*m,ua,d*_ is the excess mortality rate in municipality *m* in urban area *ua* and *département d* such as defined in Section 2.

Therefore, the time dimension does not explicitly appears, since the excess mortality rate is directly equal to the ratio between the number of deaths in April 2020 and its average over April 2019 and 2018 in each municipality. *X*_*m,ua,d*_ is a vector of controls that varies according to the specification. The most comprehensive one includes the total population and the population above 65 years old. The main coefficient of interest is *ρ* which estimates the difference across high-infection and low-infection areas of the difference in excess mortality between richer and poorer municipalities within given urban areas.

This triple-difference design allows to disentangle the impact of the epidemic from that of the lockdown that was implemented in France between March 17 and May 11. This is crucial as the lockdown effect on mortality is unclear and could be non-orthogonal to households’ income.^12^ However, the triple-difference design does not control for the risk that, coincidentally, the intensity of COVID-19 infection could be non-orthogonal to the national distribution of municipalities’ income. As displayed in Equation 1, we therefore include urban-area fixed effects (*c*_*ua*_) in our regression. Finally, we cluster standard errors at the *département* level as the most aggregated dimension of treatment definition in our design is at this level. Note that clustering at the level of urban areas does not change the nature of our results.

This model identifies the causal effect of poverty on COVID-19 related excess mortality under the sole hypothesis that, absent the COVID-19, the average difference in the evolution of mortality in April (2020 v.s. 2019 and 2018) between rich and poor cities of the same urban area would have been the same in high and low-infection *départements*.

## 4 Excess mortality due to COVID-19 and poverty

In this section, we show that, within urban areas, excess mortality due to COVID-19 was higher in poorer municipalities. We first uncover a non-linear relationship between within-urban-area income and excess mortality, showing that the poorest municipalities suffered more of COVID-19. We then turn to the estimation of Equation 1 to argue that the relationship between municipalities’ median income and excess mortality is causal.

### 4.1 A non-linear relation between income and mortality due to COVID-19

In this subsection, we show that the relation between municipalities’ income and excess mortality is not linear but driven by the poorest municipalities. These results are of importance *per se* but also because they inform our identification strategy and the way we define poor and rich municipalities.

To show that most of the excess mortality is driven by the poorest municipalities of French urban areas, we estimate Equation 2. We rank municipalities according to the median standard of living of their population, from the poorest quartile (*Q*1) to the richest quartile (*Q*4).

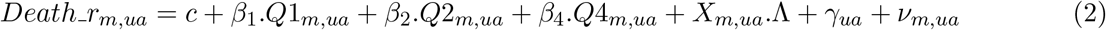

Our main dependent variable, *Death_r*_*m,ua*_ is the death ratio measured at the municipality level in April 2020 (relative to 2019 and 2018). We regress this outcome, separately for high and low-infection *départements*, on a set of dummies indicating each of the four quartiles (omitting *Q*3 as a reference category) as well as a vector *X*_*m,ua*_ of municipality characteristics and urban-area fixed effects (*γ*_*ua*_). Standard errors are clustered at the *département* level.

Figure 1 plots the estimated coefficients *β*_1_, *β*_2_ and *β*_4_ (and their 95% confidence interval) for urban areas in high-infection *départements* (panel A) and in low-infection *départements* (panel B). Figure 1(a) shows a marked difference between the excess mortality in the bottom quartile and in the other groups. On average, within the high-infection urban areas, the rise in the number of deaths that occurred in bottom-quartile municipalities (between April 2020 and both April 2019 and 2018) is 40 percentage points larger than in municipalities of the 3rd quartile. Excess mortality in the other quartiles follows a weak trend in income but no difference across these groups is significant at the 5% level. Strikingly also, such pattern is absent in low-infection urban areas. The poorest municipalities of these urban areas do not show excess mortality levels that are significantly different from richer municipalities. We take these first results as evidence that COVID-19 increased excess mortality in the poorest municipalities within high-infection urban areas. This result is robust to alternative definition of income.^13^

**Figure 1:**
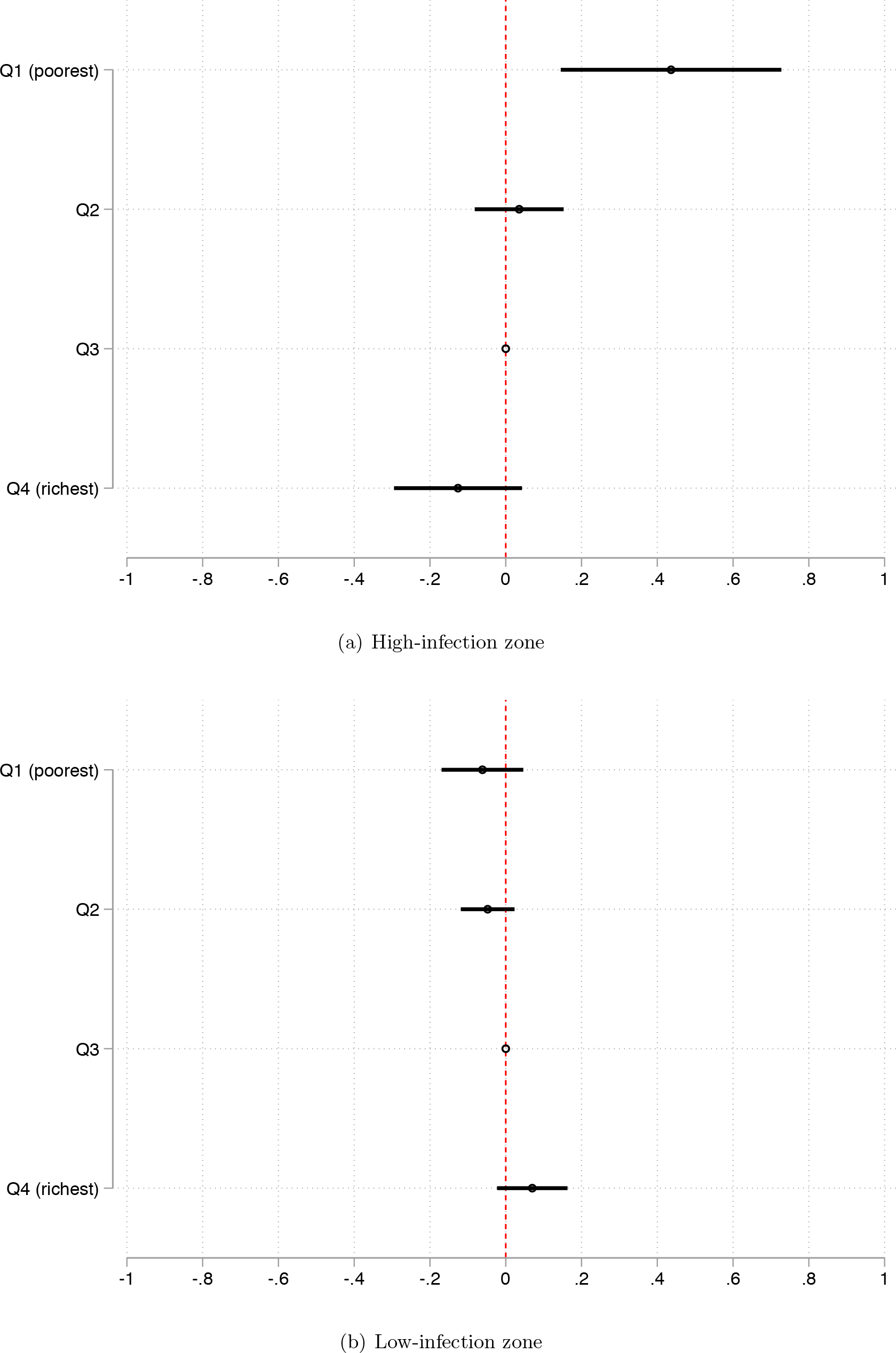
Excess mortality according to municipalities’ income. *Note:* This Figure shows the 95% confidence intervals of the coefficients estimated in Equation 2 - that is the regression of excess mortality on quartiles of the distribution of municipalities’ income, on controls and on urban areas fixed effects. Regressions are separately led in high-infection *départements* (Figure 1(a)) and low-infection ones (Figure 1(b))

### 4.2 The causal impact of poverty on excess mortality due to COVID-19

In this subsection, we argue that the relation between municipality income and excess mortality in April 2020 shown in Figure 1 is causal and due to COVID-19. Our argument is based on the main hypothesis that: absent COVID-19, the difference in the average evolution of mortality in April (2020 vs. 2019 and 2018) between richer and poorer municipalities of each urban area would be the same across (actually) high-infection and low-infection *départements*. To support this hypothesis, we first present graphical evidence of the change in mortality across municipalities’ income between high-infection and low-infection areas across time. We then estimate the corresponding triple-difference model.

Figure 2 displays the binned scatter plot of the average number of death per year, given the controls (total population, population above 65 years old) and the urban-area fixed effects. It does so separately for municipalities from the bottom quartile (red line) and the other three (blue line). The left panel is based on low-infection *départements* only. On that sample, there is a small rise in deaths in 2020 compared to previous year but the evolution is comparable across poorer and richer municipalities, although slightly higher for the lowest quartile of income. In high-infection areas however (right panel) the pattern is dramatically different: in 2020, mortality sharply increased and much more so in the lowest quartile municipalities than in the richer municipalities. Importantly, over 2018 and 2019, change in mortality was very similar in poorer and richer quartiles of these urban areas (i.e. the trends were parallel) as in low-infection *départments*. That last evidence supports the hypothesis that the evolution in urban areas would have been similar in all *départements* absent COVID-19; while it is not possible to test such hypothesis, we take Figure 2 as a strong argument in its favor.

**Figure 2:**
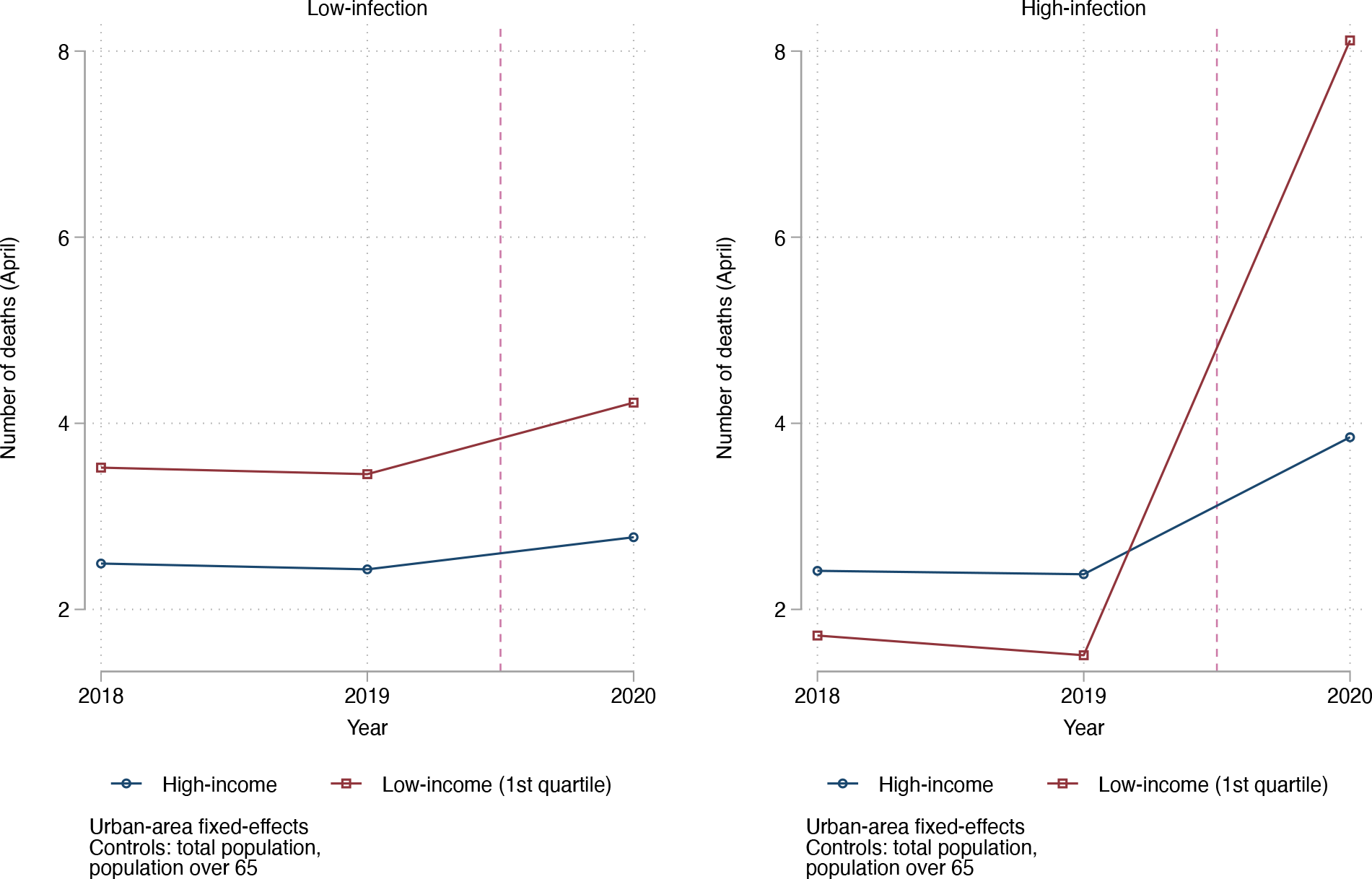
Municipalities’ mortality between 2018 and 2020 according to municipalities’ poverty level. *Note:* This Figure shows the evolution of municipalities’ average number of deaths during the month of April between 2018 and 2020, separately for municipalities whose median income is lower than the first quartile of French municipalities’ median income (red) and municipalities above that threshold (blue). The left (right) hand graph corresponds to municipalities located in low (high) infection *départements*. All estimations control for urban area fixed effects, total population and share of population above 65 years old.

Estimands of Equation 1 are displayed in Table 1. They prove that the relation between poverty and excess mortality due to COVID-19 is positive and that it is driven by the poorest municipalities. Estimands from our preferred specification are displayed in panel A of columns (1) and (2). In line with what precedes, the poverty cut-off is the first quartile of municipalities’ median income. As a robustness check, we also show the result of a similar estimation where the cut-off is the median income (see columns (3) and (4)).

**Table 1:** Excess mortality due to COVID-19 according to municipalities’ poverty level.

|  | (1) | (2) | (3) | (4) |
| --- | --- | --- | --- | --- |
| Infected Region | 0.56765***<br>(0.15339) | 0.50295***<br>(0.14053) | 0.55603***<br>(0.15971) | 0.49125***<br>(0.14508) |
| Very poor (Quart1) | -0.03873<br>(0.04827) | -0.04777<br>(0.04885) |  |  |
| Infected*Quart1 | 0.50212***<br>(0.13412) | 0.42904***<br>(0.11738) |  |  |
| Poor |  |  | -0.05926*<br>(0.03207) | -0.05818*<br>(0.03177) |
| Infected*Poor |  |  | 0.27501***<br>(0.07986) | 0.21982***<br>(0.06735) |
| Control outcome mean | 1.105 | 1.105 | 1.130 | 1.130 |
| Urban areas FE | ✓ | ✓ | ✓ | ✓ |
| Controls | . | ✓ | . | ✓ |

Our preferred specification indicates that, in municipalities above the cut-off income in low-infection *départements*, the average number of deaths in April 2020 is 110% of the average number of fatalities in 2019 and 2018. This limited rise is potentially due to COVID-19 but we do not disentangle its causes and consider it as a baseline scenario for 2020 (that is the most conservative approach). The contrast with high-infection areas is striking: there, mortality in April 2020 is 160% of that of April 2019 and 2018 (i.e. a 0.50 increase in the average excess mortality of 1.1 in non-poor, low-infection municipalities). Most importantly for us, while the impact of income on excess mortality is not significantly different from zero in low-infection *départements*, it is strongly and significantly positive in high-infection *départements*. In each urban area of these *départements*, COVID-19 has caused an excess mortality that is larger by 38 (43-5) percentage points on average in municipalities of the bottom quartile of the national distribution of median incomes than in richer municipalities. That is to say, poorest municipalities saw their mortality double.

The nature of these results does not change if we take the median of municipalities’ income as a cut-off (see columns (3) and (4)).

## 5 Potential mechanisms

In this section, we explore the role of two potential mechanisms that could explain the relationship between poverty and COVID-19 related excess mortality. Poverty is multidimensional and we do not aim to identify all the potential channels of transmission from poverty to excess mortality. We rather focus on two specific factors we are able to observe with high-quality data. In subsection 5.1, we explain what dimensions we chose to explore. We next turn to present mechanism-specific results in subsections 5.2 to 5.4 before trying to quantify their respective role in subsection 5.5.

### 5.1 Two Potential Channels

Informed by previous literature and the current knowledge of how COVID-19 is transmitted, we decided to focus on (i) labor-market related measures of exposure to the virus and (ii) housing conditions as two potential channels between poverty and excess mortality. We of course acknowledge that these are only two potential mechanisms among many, and we notably ignore the role of comorbidities known to be related with poverty and that most likely played a role in the observed phenomenon (Wiemers et al., 2020; Raifman and Raifman, 2020).

Regarding labor-market conditions, we explore the idea that low-paid occupations are also more exposed to COVID-19 transmission. On this aspect, we take into account the effect of the lockdown by contrasting the role of occupational exposure and the specific role of continuing sectors. The main hypothesis behind this mechanism is that low-paid occupations are both more present in poorer municipalities and more exposed to COVID-19 transmission. To confirm this link, we first build a measure of exposed occupations using the DEFIS database: for each municipality, we compute an “index of frequent contact” based on the occupation of all employed residents. We next turn to identifying the sectors that continued during lockdown using an official decree. See section 2.3 for more details on these two measures. Table 2 shows the strength of the link between poverty and our labor-market-related measures, once included our baseline covariates and urban-area fixed effects. In both cases,, municipalities of the poorest quartile have more exposed occupations (first column of Panel A) and more workers in continuing sectors (second column of Panel A).^14 15^

**Table 2:** Association between poverty and occupation and housing variables.

|  | Index of frequent contact | Rate of authorized jobs | Share of over-crowded housing |
| --- | --- | --- | --- |
| *Panel A: All urban areas* |  |  |  |
| Main coefficient | 0.48390***<br>(0.16111) | 0.25848***<br>(0.09399) | 2.46026***<br>(0.25582) |
| Control outcome mean | 0.092 | 1.038 | 1.042 |
| Observations | 14684 | 14681 | 14360 |
|  | Share of multi-generational<br>hh with at least one worker |  |  |
| *Panel B: Urban areas with more than 2000 inhabitant cities* |  |  |  |
| Main coefficient | 2.03794**<br>(0.79269) |  |  |
| Control outcome mean | 1.314 |  |  |
| Observations | 3741 |  |  |
| Urban areas FE | ✓ | ✓ | ✓ |
| Controls | ✓ | ✓ | ✓ |

\* $p < 0.05$ , \*\* $p < 0.01$ , \*\*\* $p < 0.001$ . Standard errors in parentheses are clustered at the *département* level. Each column reports the result of a separate regression of poverty on each mechanism variable. Poor is defined as belonging to the first quartile of the median income distribution. Controls include total population and population over 65 years old. The control outcome mean line reports the mean of the excess mortality rate in low-infection municipalities.

Regarding housing conditions, we mainly employ a demographic measure from the Census data: the share of overcrowded housing units in the municipality (see 2.4). By multiplying the probability of social contacts (either within the household or by increasing the need of going out), especially in closed spaces, overcrowding is an obvious candidate for COVID-19 transmission. In Table 2, column 4 of Panel A indicates that within urban areas and given population size and age-structure, increasing the share of overcrowded housing units increases the probability of a municipality to fall in the poorest quartile.^16^ In other words, poorest municipalities of urban areas indeed have more overcrowded housing units.

We complement our study of labor-market condition housing demographics by taking one measure at the interaction of these two dimensions. We compute the share of municipalities’ households that gather at least one member aged 65 or more and one member from a younger generation who is currently employed (see section 2.5 for more details on this measure). This measure requires occupation information from the Census, which are only measured in municipalities with at least 2,000 inhabitants and we thus report the associated results in Panel B of Table 2. Here again, poorest municipalities are much more likely to have such type of multigenerational households.^17^ The correlation between the share of such households and the probability for a municipality to belong the poorest quartile is positively and highly significant.

### 5.2 Labor Market and Exposure to COVID-19

In this section, we re-estimate the triple difference model exposed in Equation 1 but substituting labor-market-related measures in place of the poverty indicator. The results are presented in the first two columns of Panel A of Table 3. The Table only reports the main coefficient of interest (i.e. the *ρ* of Equation 1) which interpretation remains: it is the estimated difference (across high-infection and low-infection areas) of the difference in excess mortality due to the increase of one unit of the main independent variable within each urban area. In other words, the main coefficient indicates the magnitude of excess mortality, within one municipality, due to having more of a specific type of labor-market conditions.

**Table 3:** Effect of Covid-19 on excess mortality - All mechanisms.

|  | Occupational exposure mechanism |  | Housing mechanism |
| --- | --- | --- | --- |
|  | Index of frequent contact | Rate of authorized jobs | Share of over-crowded housing |
| *Panel A: All urban areas* |  |  |  |
| Main coefficient | 2.56144***<br>(0.81134) | -0.60416<br>(0.47456) | 5.65638***<br>(1.02754) |
| Control outcome mean | 1.314 | 1.314 | 1.319 |
| Observations | 13471 | 13455 | 12918 |
| *Panel B: Urban areas with more than 2000 inhabitant cities* |  |  |  |
| Main coefficient | 3.95791<br>(2.75105) | 1.15564<br>(2.85059) | 5.84591***<br>(1.93072) |
| Outcome mean | 1.556 | 1.556 | 1.562 |
| Observations | 3671 | 3671 | 3653 |
| Interaction of both mechanisms |  |  |  |
| Share of multi-generational hh with at least one worker |  |  |  |
| *Panel B: Urban areas with more than 2000 inhabitant cities* |  |  |  |
| Main coefficient | 12.16877**<br>(5.82533) |  |  |
| Control outcome mean | 1.314 |  |  |
| Observations | 3671 |  |  |
| Urban areas FE | ✓ | ✓ | ✓ |
| Controls | ✓ | ✓ | ✓ |

Only one of our two labor-market indices shows up significantly: the index of frequent contact. For that variable (first column of Panel A of Table 3), our favorite estimate indicates that when the share of employed inhabitants frequently exposed to other individuals while working goes from 0 to 100% in highly infected *départments*, excess mortality in April increases by 2.5, i.e. more than twice the average excess mortality in our sample. In other words, municipalities of high-infection *départements* where all employed inhabitants are exposed would have recorded 2.5 times more deaths in April 2020 compared to the average number of death in April 2018 and 2019. Put differently, increasing the share of exposed workers by one standard-deviation (0.046 in both high- and low-infection urban areas) increases predicted excess mortality by 0.12. This is an important effect. Continuing-sector effect on the other hand appear to be insignificant. There are three main potential explanations: first, it could be that we measure a true null effect of lockdown whereby COVID-19 did not spread more in these continuing industries. Second, it could be that death toll of April are only partially affected by lockdown (given the delays between infection and death). Third and finally, measurement errors could affect the definition of continuing industries as our measure from the decree actually is closer to an intention-to-treat than an actual treatment status.

### 5.3 Housing Conditions

In this section, we re-estimate the triple difference model exposed in 1 but substituting housing-condition measures in place of the poverty indicator. The results are presented in the last column of Panel A of Table 3. Reported coefficient is still the equivalent of the *ρ* of Equation 1 but taking housing conditions variables instead of our measure of poverty.

Our measure is the municipality-level share of housing units that are overcrowded. As shown by Table 3, housing overcrowding indeed caused an increase in observed excess mortality: the coefficient is positive and highly significant. The point estimate suggests that increasing the share of overcrowded housing units from 0 to 100% increases excess mortality by 5.6 points. To get a sense of this effect, increasing the share of over-crowded housing units by one standard-deviation (.025) increases the excess mortality by 14 percentage points. This is a very important effect, that is comparable to the estimates related to contacts on the labor market.

### 5.4 Interacting Labor-Market and Housing Conditions

In Panel B, we estimate the effect of the share of households including both elderly and employed individuals: here as well, that demographic structure seems to foster the transmission of COVID-19 and its associated mortality. We find that increasing the share of such household from 0 to 100% would increase excess mortality by 12. Although this coefficient is very high, such households only represent 2% of the residents on average. An increase of the share of multi-generational households by one standard-deviation (0.01) yields an increase in excess mortality of 12 percentage points. Since this last measure is only observed on a sample of municipalities with more than 2,000 inhabitants, we also reproduced our results of Panel A (for our main three variables) on that sample. All the point estimates are higher on this sample, but so are standard errors, leading to non-significant results in two out of three cases.

### 5.5 Disentangling the role of each mechanism

Understanding which mechanism prevails is crucial for the design of the best policy response. Therefore, we perform a horse race between variables related to occupational exposure and housing conditions. Using the same triple-difference setting as in the main regression, Table 4 regresses the excess mortality rate on poverty alone, and on poverty with each mechanism variable separately. Finally, the last column includes both poverty and all the mechanism variables. All covariates have been normalized to ease comparison. When comparing column (1) with columns (2) to (4), we observe that the inclusion of the share of overcrowded housing makes the coefficient of poverty shrink. Other variables have only a minor effect on the poverty coefficient. In the last column, poverty, housing conditions, and the index of occupational contact still play a significant role in explaining excess mortality. The initial coefficient of poverty diminishes by 38% when all covariates are included. All in all, it suggests that both the housing conditions and the frequency of social contacts through occupations are important determinants of the relatively higher excess mortality due to COVID-19 in poor municipalities. However, housing conditions appear to be the main determinant.

**Table 4:** Effect of each covariate on excess mortality due to COVID-19.

|  | Excess Mortality Rate, April 2020 |  |  |  |  |
| --- | --- | --- | --- | --- | --- |
| Infected Region=1 × Very poor (Quart1) | 0.17657***<br>(0.04831) | 0.16974***<br>(0.04423) | 0.17918***<br>(0.04820) | 0.10817**<br>(0.04338) | 0.11021**<br>(0.04255) |
| Infected Region=1 × Index of frequent contact | | 0.12227***<br>(0.04005) | | | 0.08902**<br>(0.03998) |
| Infected Region=1 × Rate of authorized jobs | | | -0.06029*<br>(0.03462) | | -0.05202<br>(0.03505) |
| Infected Region=1 × Share of over occupied housing units | | | | 0.12320***<br>(0.02768) | 0.11429***<br>(0.02855) |
| Urban areas FE | ✓ | ✓ | ✓ | ✓ | ✓ |
| Controls | ✓ | ✓ | ✓ | ✓ | ✓ |
| Control outcome mean | 1.038 | 1.038 | 1.038 | 1.043 | 1.043 |
| Adjuster $R^2$ | 0.0441 | 0.0489 | 0.0454 | 0.0527 | 0.0570 |
| Observations | 13219 | 13219 | 13216 | 12912 | 12910 |

\* $p < 0.05$ , \*\* $p < 0.01$ , \*\*\* $p < 0.001$ . Standard errors in parentheses. This table shows the result of regressing municipalities' excess mortality in April 2020 on a dummy indicating that the municipality is located in a high-infection *département*, a variable measuring either poverty, housing conditions or occupational exposure and the interaction of these variables, using the ratio definition of excess mortality (i.e. number of deaths in April 2020 as compared to April 2018 and 2019). The first columns only examines the poverty channel. Columns (2) to (4) respectively include one additional variable capturing either the occupation or housing mechanism. The last column includes both the poverty dummy and all the mechanism variables. All regressions include urban areas fixed-effects and control for total population and for the age structure in the municipality. Each covariate has been normalized. The control outcome mean line reports the mean of the excess mortality rate in low-infection municipalities.

This analysis in confirmed by a Oaxaca-Blinder decomposition that quantifies the share of the gap in excess mortality between poor and rich municipalities within urban areas in high-infection *départements* that is explained by the included covariates (Oaxaca, 1973; Blinder, 1973). The top part of Table A1 of Appendix 2 reports the excess mortality rate in poor and non-poor municipalities in high-infection *départements*, as well as the share of the difference between both rates that is explained by the included variables, and the share that is unexplained. The middle and bottom parts of the table describe, respectively, the contribution of each variable to the explained and unexplained shares of the difference. The first column of Table A1 indicates that 63% of the 39 percentage points difference in excess mortality rate between rich and poor municipalities within urban area is explained by the variables capturing occupational exposure and housing conditions. The sole share of overcrowded housing explains virtually all of these 63%. The inclusion of population controls in column (2) makes the explained share jump to 91% of the total difference, but does not affect much the respective weight of each covariate. Although this decomposition is based on certain assumptions, it confirms that housing conditions are the main determinant of the within-urban-area gap in excess mortality between rich and poor municipalities. They account for 46% to 64% of the total difference.

## 6 Robustness checks

### 6.1 Excess mortality according to municipality’s income by age group

Age is one of the main determinants of COVID-19 mortality (Wu and McGoogan, 2020): the National Public Health Agency, in its report of March 24, estimates the average age at death due to COVID-19 at 81.2. Therefore, we expect most of the variation in excess mortality to come from the top of the age distribution. Figure A2 of Appendix 3 shows the coefficient from the same triple-difference specification as in Table 1, for each age group separately. The coefficient for the total population is reproduced at the left hand side of Figure A2. We indeed observe a pattern increasing with age: among people over 85 y.o., the difference in excess mortality between rich and poor municipalities within a given urban area is larger by 63 percentage points in high-infection *départments* than in low-infection ones. If the coefficients in the four top age groups are not significantly different from each other, they are significantly different from the coefficients for the population below 50 y.o. That supports our previous findings by showing that they are consistent with the empirical fact that most of COVID-19 related mortality is to be found among the elderly. It also indicates that the causal impact of poverty on COVID-19 related mortality is particularly strong for this age group.

### 6.2 Alternative excess mortality measure

Our preferred excess mortality measure described in Section 2 allows to account for population size, and is in line with the recommendations of the National Public Health Institute. However, we reproduce the main specification of Table 1 on the number of deaths rather than on the excess mortality rate. Table A2 of Appendix 3 shows the triple-difference coefficients of a regression of the death toll in April on three dimensions (and their interactions): (i) living in a high-v.s. low-infection region; (ii) living in a poor v.s. rich municipality, (iii) in 2020 v.s. the average between 2019 and 2018. It indicates that, within urban areas in high-infection *départements*, being a poor municipality (i.e. belonging to the first quartile of the income distribution) increases the number of deaths in April 2020 by 9 units relative to a baseline number of deaths in rich municipalities during the pre-COVID period of 3.8. In low-infection *départements*, the average pre-COVID difference in death toll between rich and poor municipalities of the same urban area was about 1.6 units. It has only increased by 0.7 units after COVID-19. The main effect slightly decreases if we define poverty relative to the median of the income distribution, but remains in the same order of magnitude. Results are therefore consistent with our main specification, and are not sensitive to the definition of the outcome variable.

## 7 Conclusion

In this paper, we use administrative data to investigate the role of household income in explaining the heterogeneity of excess mortality due to COVID-19 across French municipalities, a very small administrative unit. In France, the lock-down was uniformly implemented over the French territory between March 17 and May 11. As a consequence, it froze municipalities at very different stages of diffusion of the disease. We exploit this quasi-experimental setting to implement a triple-difference design comparing the mortality over time, across high- and low-infection local areas (*départements*) and across the distribution of municipalities’ income. This triple-difference allows us to estimate the causal impact of poverty on COVID-19 related mortality, net of any lockdown effect. We show that, in municipalities located in one of the high-infection *départements*, COVID-19 has caused a 50 percentage point rise in the number of deaths in April 2020 on average in comparison with the average number of deaths in April 2018 and April 2019. Among these *départements*, municipalities that are part of the bottom quartile of the national distribution of municipalities’ income saw an increase in mortality of around 100%, about 38 percentage points higher than in richer high-infection municipalities. Given this first set of results, we investigate the role of two potential mechanisms related to labor-market and housing conditions. We find that poorer municipalities have both a higher share of workers frequently in contact with the public and of over-occupied housing. Both these dimensions explain a significant part of the link between poverty and COVID-19 related mortality. Our analysis concludes that housing conditions are the main determinant among these two.

## Data Availability

Most data used in this paper is publicly available. The rest of the data is available for research purpose under agreement with the CASD.

## A Appendix 1

*by alphabetical order*

### Data sources

**DADS** (for “Déclarations Annuelles des Données Sociales”) is a matched employer-employee exhaustive data set that covers all employees in private and public sectors (outside of agriculture) in the year 2016. The DADS is an administrative data set that all employers must report yearly to social security authorities and tax administration. The version available to researchers is provided by the INSEE. In particular, DADS contain information about all positions occupied by any worker in a specific year, with start and end date at the daily level. In particular, for each position held by one individual we observe: the occupation, the firm and sector, both location of work and residency and the number of hours spent. When using DADS to build municipality-level indicators we rely on the municpality of residency.

**Daily Deaths Files** (“fichier des décès quotidiens”) from INSEE record each death that occurred between January 1st, 2018 and June 5, 2020. For each death, we observe information relative to the event (the date and municipality of death as well as a category about the place of death - hospital or clinic, home, care home, etc.) and the individual (*département* of residency, sex, date of birth). INSEE made these files available at a higher frequency during the covid-19 crisis, but this comes at the expense of some quality checks. The files are originally recorded by municipalities and then gradually added to the INSEE data sets, one cannot exclude that the files were incomplete by the time of our analysis although we updated them frequently (last update: June 30^*th*^).

URL: https://www.insee.fr/fr/statistiques/4487854

***DEFIS*** (“Dispositif d’enquêtes sur les formations et itinéraires des salariés”) is a database produced by the Céreq. It comes from a survey led on 4,500 firms and 16,000 of their workers as of 2013. Firms are representative of the whole private sector for firms with more than 10 workers but only of some sub-sectors for smaller firms. In this paper, we use the survey led on workers in 2015. The question of interest relates to contacts with the public in the course of their work. It refers to the current job of workers who have not changed firm since 2013 and to the one of 2013 for workers who changed firm since then. URL: https://www.cereq.fr/en/data-access-lifelong-learning-and-vocational-training-surveys-defis-cvts-base-redefis-employee https://www.overleaf.com/project/5eea2511dc82cf0001a68a74

***Filosofi* database** (a French acronym for “Localised disposable income system”), 2014 version, contains a series of local measures on income and poverty. INSEE uses fiscal and social benefits data to compute these indicators at the municipality level. In particular, we are interested in the median income of each municipality’s inhabitants. URL: https://www.insee.fr/fr/metadonnees/source/operation/s1451/presentation

**Population Censuses** are used to compute a series of measures at the municipality level. Each year: (i) a fifth of municipalities with less than 10,000 inhabitants are covered by an exhaustive census and (ii) a sample of 8% of the population of municipalities with 10,000 or more inhabitants are surveyed. In-depth questions relative to economic activity and family structure of households are only available for about 20% of the households of municipalities under 10,000 inhabitants and about 40% of the households in the other municipalities (10,000 inhabitants or more). Analyses of in-depth questions cannot be done for municipalities with less than 2,000 inhabitants as samples get too small. We use INSEE annual population counts and both exhaustive and partial census data files.

URL: https://www.insee.fr/fr/information/2383265

**Urban Areas** are defined by the INSEE as a group of neighboring municipalities encompassing an urban centre and its periphery made of municipalities where at least 40% of employed inhabitants works in the urban centre, as of 2010. We often consider only the big urban areas, where the centre contained 10,000 jobs or more as of 2010. Big urban areas represent 80% of the French mainland population in 2014.

## B Appendix 2

### Additional Tables and Figures

#### 1 Institutional framework

**Figure A1:**
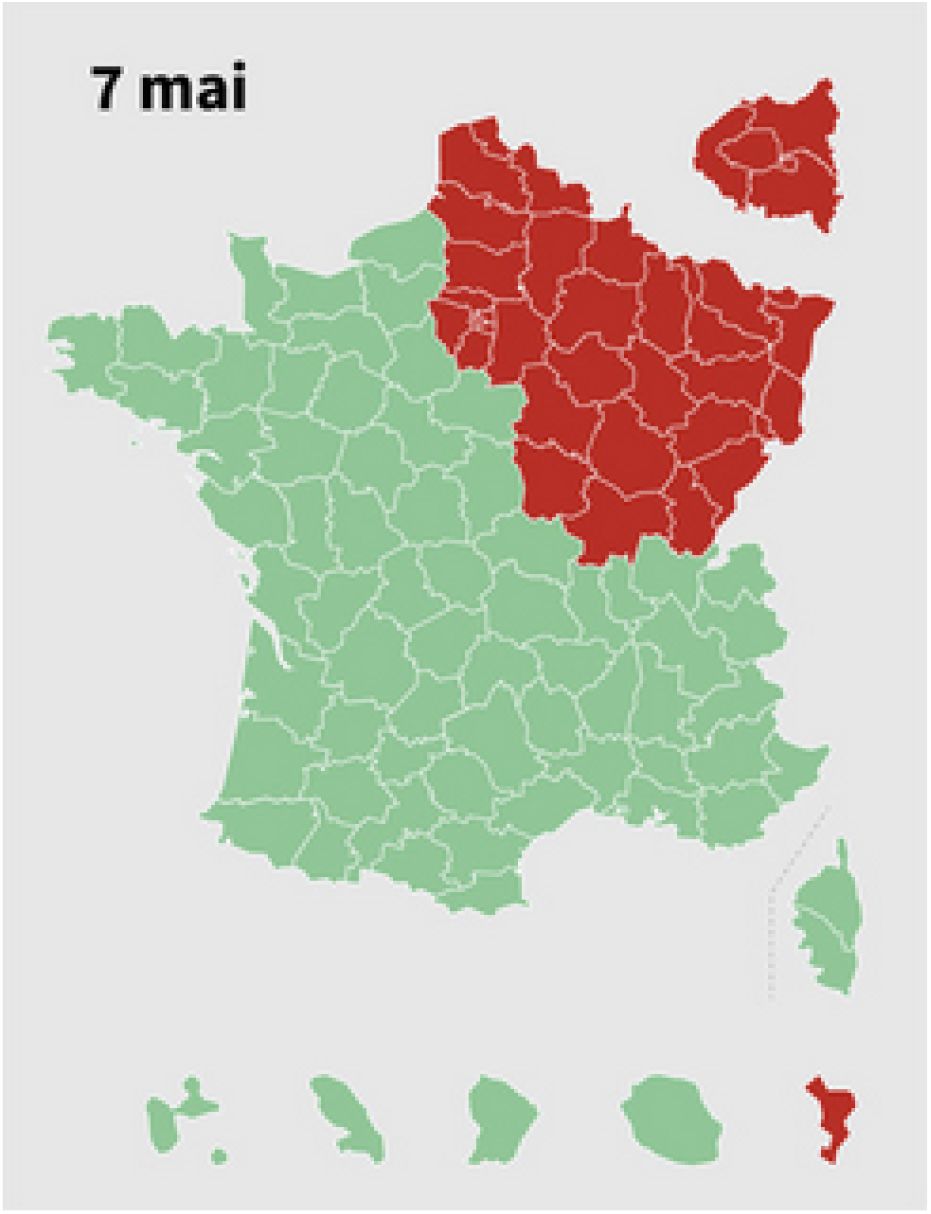
High-(red) and low-infection (green) *Départements* as of May 7, 2020. *Note:* Map of *Départements* issued by the Government on May 7, 2020. See source.

#### 2 Oaxaca-Blinder decomposition

**Table A1:** Oaxaca-Blinder decomposition of the excess mortality gap between poor and rich municipalities in high-infection areas.

|  | Excess Mortality Rate, April 2020 |  |
| --- | --- | --- |
| Very poor (Quart1) = 0 | 1.20146***<br>(0.02016) | 1.20146***<br>(0.02017) |
| Very poor (Quart1) = 1 | 1.59473***<br>(0.06071) | 1.59473***<br>(0.06091) |
| Difference | -0.39327***<br>(0.06397) | -0.39327***<br>(0.06416) |
| Explained | -0.24753***<br>(0.03606) | -0.35915***<br>(0.04512) |
| Unexplained | -0.14574**<br>(0.06488) | -0.03412<br>(0.07064) |
| *Explained* |  |  |
| Share of over occupied housing units | -0.25050***<br>(0.03110) | -0.18111***<br>(0.02929) |
| Index of frequent contact | -0.00523<br>(0.00421) | -0.00363<br>(0.00309) |
| Rate of authorized jobs | 0.00989**<br>(0.00402) | 0.00897**<br>(0.00382) |
| Urban Area, code | -0.00168<br>(0.01134) | -0.00162<br>(0.01129) |
| Population Municipality, 2014 |  | 0.62475***<br>(0.15872) |
| Pop. 65-74 |  | -1.19171***<br>(0.24785) |
| Pop. 75-84 |  | 0.08588<br>(0.20125) |
| Pop. 85+ |  | 0.29932***<br>(0.10717) |
| *Unexplained* |  |  |
| Share of over occupied housing units | 0.04933<br>(0.03896) | -0.03182<br>(0.04847) |
| Index of frequent contact | -0.00217<br>(0.00478) | -0.00359<br>(0.00523) |
| Rate of authorized jobs | -0.00139<br>(0.02639) | -0.00447<br>(0.02646) |
| Urban Area, code | -0.01505<br>(0.05237) | -0.02028<br>(0.05325) |
| Population Municipality, 2014 |  | -0.60932**<br>(0.25399) |
| Pop. 65-74 |  | 1.28955**<br>(0.50690) |
| Pop. 75-84 |  | -0.16959<br>(0.46324) |
| Pop. 85+ |  | -0.26405<br>(0.23183) |
| Constant | -0.17646**<br>(0.08868) | -0.22055**<br>(0.10048) |
| Observations | 5339 | 5339 |

\* $p < 0.05$ , \*\* $p < 0.01$ , \*\*\* $p < 0.001$ . Standard errors in parentheses. This table reports the output of a Oaxaca-Blinder decomposition (Oaxaca, 1973; Blinder, 1973) of the gap in excess mortality between rich and poor municipalities in high-infection areas. It indicates that the share of this gap that is explained by included covariates. Each covariate has been normalized.

#### 3 Robustness checks

##### Excess mortality according to municipality’s income by age group

**Figure A2:**
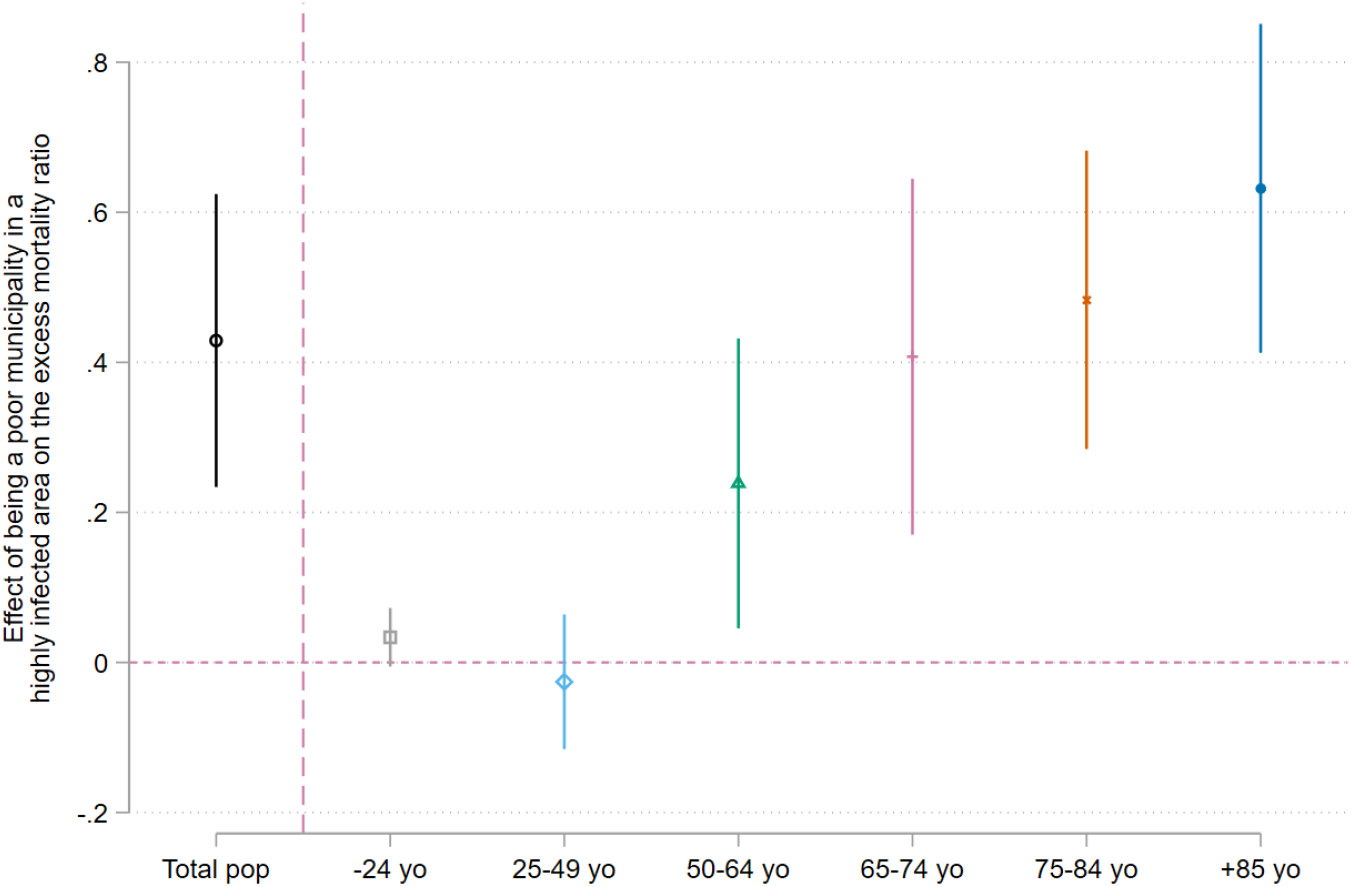
Differential effect of Covid-19 in rich and poor municipalities by age group. *Note:* This graph plots the triple difference coefficient for each age group separately. It measures the effect of the interaction term between the high-infection region dummy and the poverty dummy on the excess mortality in April for each age group.

##### Alternative excess mortality measure

**Table A2:** Death toll in April according to municipalities’ median income.

|  | Death toll, April |  |  |  |
| --- | --- | --- | --- | --- |
| Infected Region=1 | 4.44356**<br>(1.88259) | 1.47092<br>(0.95593) | 4.43288**<br>(1.92102) | 1.43097<br>(0.97176) |
| Very poor (Quart1)=1 | 4.56109***<br>(0.80568) | 1.61709**<br>(0.79419) |  |  |
| Infected Region=1 × Very poor (Quart1)=1 | 3.50447**<br>(1.34682) | 0.86855<br>(0.65567) |  |  |
| 2020=1 | 0.31417***<br>(0.08710) | 0.31417***<br>(0.08710) | 0.30775***<br>(0.08374) | 0.30775***<br>(0.08374) |
| Infected Region=1 × 2020=1 | 1.13961***<br>(0.42557) | 1.13961***<br>(0.42559) | 1.14070**<br>(0.43813) | 1.14070**<br>(0.43815) |
| Very poor (Quart1)=1 × 2020=1 | 0.41920*<br>(0.24234) | 0.41920*<br>(0.24235) |  |  |
| Infected Region=1 × Very poor (Quart1)=1 × 2020=1 | 4.62875**<br>(1.97916) | 4.62875**<br>(1.97925) |  |  |
| Poor=1 |  |  | 2.75368***<br>(0.50238) | 0.99277*<br>(0.51641) |
| Infected Region=1 × Poor=1 |  |  | 2.02596**<br>(0.93545) | 0.61896*<br>(0.32422) |
| Poor=1 × 2020=1 |  |  | 0.12968<br>(0.08445) | 0.12968<br>(0.08446) |
| Infected Region=1 × Poor=1 × 2020=1 |  |  | 1.68545**<br>(0.82111) | 1.68545**<br>(0.82115) |
| Urban areas FE | ✓ | ✓ | ✓ | ✓ |
| Controls | . | ✓ | . | ✓ |
| Control outcome mean | 2.329 | 2.329 | 1.981 | 1.981 |
| Observations | 44457 | 44457 | 44457 | 44457 |

\* $p < 0.05$ , \*\* $p < 0.01$ , \*\*\* $p < 0.001$ . Standard errors in parentheses clustered at the *département* level. This table shows the result of regressing municipalities' death toll in April on a dummy indicating that the municipality is located in a high-infection region (*département*), a dummy indicating that the municipality's median income is under the poverty cut-off, a dummy *Post* equal to one in 2020, and the interaction of these dummies. The poverty cut-off in columns (1) and (2) corresponds to the first quartile of municipalities' median income, while it corresponds to the median of municipalities' median income in columns (3) and (4). All regressions include urban areas fixed-effects. Regressions in columns (2) and (4) further control for total population and for the age structure in the municipality.

## Footnotes

1 Source: European Centre for Disease Prevention and Control (https://www.ecdc.europa.eu/en/geographical-distribution-2019-ncov-cases).

2 In particular, measures based on confirmed cases are likely to introduce a substantial downward bias due to limited (and unequally distributed) testing capacities (Borjas, 2020; Roser et al., 2020). For example, projections by Silverman et al. (2020) suggest that almost 80 percent cases of COVID-19 in the US were never diagnosed. Scarpetta et al. (2020) further show that France ranks among the worst OECD countries with respect to the number of tests per inhabitants.

3 More specifically, differential effects of lockdown and/or seasonal diseases between rich and poor municipalities would bias our estimation of the heterogeneous impact of COVID-19 on mortality across municipalities.

4 Government records indicate that, on March 17, the intensive care units occupancy rate was 8% in low-infection *départements* vs. 34.5% in high-infection *départements*.

5 Among others, further mechanisms may also include: greater levels of air pollution in poorer areas (Cole et al., 2020; Persico and Johnson, 2020) and lower levels of compliance to lockdown and to self-protection measures among low-income individuals Papageorge et al. (2020), among others. While population density is also an often debated mechanism, we do not treat it as a potential mechanism given its negative correlation with poverty in our data.

6 For a discussion on the quality of the data, see the information provided by INSEE. We follow the guidelines detailed in this blog post.

7 We use the average of years 2018 and 2019 because taking several years makes our measure less sensitive to random events occurring in a given year. For data availability reasons, we cannot go further back in time, since death data before 2018 do not include, at the moment, information on the municipality of residency. However, INSEE advises against using too many years because demographic change makes the comparison of the number of deaths less relevant as we go further back in time.

8 However, we systematically control for total population and population over 65 years old in our regressions.

9 In its report of March 24, the National Public Health Agency (*Santé Publique France*) explains that the number of deaths of people under 65 y.o. without pre-existing conditions accounts for 2.4% of total deaths due to COVID-19 in France.

10 Relative to municipalities, *départements* are higher administrative units. There are 94 *départements* in mainland France, excluding Corsica.

11 Workers are localised according to their municipality of residency and regardless of their municipality of work.

12 On the impact of the lockdown on mortality rates see Brodeur et al. (2020) - who have shown that deaths related to traffic accidents have dropped - or Bradbury-Jones and Isham (2020) - on the rise of domestic violence.

13 For instance, we also partitioned the distribution of incomes by deciles. In the high-infection areas, the bottom three deciles show similar excess mortality levels that are significantly different from municipalities in the 5^*th*^ decile. Deciles four as well as deciles six to ten are not significantly different in terms of mortality from the 5^*th*^ decile. Hence our choice to work at the quartile level in the end.

14 In Panel A of the Tables, the outcome means indicates that 9% of municipalities in our sample belong to the first quartile (that share is unsurprisingly lower than 25% since our sample is made of within-urban areas municipalities, which are on average richer than rural municipalities).

15 The first column indicates that, within an urban area, going from virtually 0 to 100% of exposed workers increases the probability to fall in the poorest quartile by 48 percentage points. Or, a one standard deviation increase in the share of exposed workers is associated with an increase in excess mortality of 2.3 percentage points. The second column shows that going from 0 to a 100% of inhabitants working in a continuing sector increases the probability to fall in the poorest quartile by 26 percentage points. This is also equivalent to an increase of 1.5 percentage points when the share of authorized job increases by one standard deviation.

16 The coefficient associated to over-crowed housing units indicates that going from 0 to 10% of over-crowded housing units in a municipality is associated with an increase of excess mortality of 24 percentage points. A one standard-deviation increase in over-crowded housing units would be associated with a 6 percentage points increase in excess mortality.

17 The coefficient indicates that going from a 0 to 100% of households with both one member older than 65 and a worker increases the probability to be in the poorest quartile by 203 percentage point. That is, an increase of excess mortality of 2.4 percentage points when the share of multi-generational households increases by one standard deviation.

## References

Abi Adams-Prassl, Teodora Boneva, Marta Golin, and Christopher Rauh. Inequality in the impact of the coronavirus shock: Evidence from real time surveys. 2020.

Nancy E Adler and David H Rehkopf. Us disparities in health: descriptions, causes, and mechanisms. Annu. Rev. Public Health, 29:235–252, 2008.

Milena Almagro and Angelo Orane-Hutchinson. The determinants of the differential exposure to covid-19 in new york city and their evolution over time. Covid Economics: Vetted and Real-Time Papers, (13), 2020.

Annette Alstadsæter, Bernt Bratsberg, Gaute Eielsen, Wojciech Kopczuk, Simen Markussen, Oddbjorn Raaum, and Knut Røed. The first weeks of the coronavirus crisis: Who got hit, when and why? evidence from norway. Technical report, National Bureau of Economic Research, 2020.

Andrea Ascani, Alessandra Faggian, Sandro Montresor, et al. The geography of covid-19 and the structure of local economies: the case of italy. Technical report, 2020.

Alan S. Blinder. Wage Discrimination: Reduced Form and Structural Estimates. The Journal of Human Resources, 8(4):436–455, 1973. ISSN 0022-166X. doi: 10.2307/144855. URL https://www.jstor.org/stable/144855. Publisher: [University of Wisconsin Press, Board of Regents of the University of Wisconsin System].

George J. Borjas. Demographic Determinants of Testing Incidence and COVID-19 Infections in New York City Neighborhoods. SSRN Scholarly Paper ID 3572329, Social Science Research Network, Rochester, NY, April 2020. URL https://papers.ssrn.com/abstract=3572329.

George J Borjas and Hugh Cassidy. The adverse effect of the covid-19 labor market shock on immigrant employment. Technical report, National Bureau of Economic Research, 2020.

Caroline Bradbury-Jones and Louise Isham. The pandemic paradox: The consequences of COVID-19 on domestic violence. Journal of Clinical Nursing, 29(13-14):2047–2049, 2020. ISSN 1365-2702. doi: 10.1111/jocn.15296. URL https://onlinelibrary.wiley.com/doi/abs/10.1111/jocn.15296.eprint: https://onlinelibrary.wiley.com/doi/pdf/10.1111/jocn.15296.

Abel Brodeur, Nikolai Cook, and Taylor Wright. On the Effects of COVID-19 Safer-At-Home Policies on Social Distancing, Car Crashes and Pollution. Technical Report 13255, Institute of Labor Economics (IZA), May 2020. URL https://ideas.repec.org/p/iza/izadps/dp13255.html. Publication Title: IZA Discussion Papers.

Matthew Cole, Ceren Ozgen, and Eric Strobl. Air pollution exposure and covid-19. 2020.

Dhaval M Dave, Andrew I Friedson, Kyutaro Matsuzawa, Joseph J Sabia, and Samuel Safford. Black lives matter protests, social distancing, and covid-19. Technical report, National Bureau of Economic Research, 2020.

Robert W Fairlie, Kenneth Couch, and Huanan Xu. The impacts of covid-19 on minority unemployment: First evidence from april 2020 cps microdata. Technical report, National Bureau of Economic Research, 2020.

Sté phane Foucart and Sté phane Horel. Dé pistage du coronavirus : les raisons du fiasco français sur les tests. Le Monde.fr, April 2020. URL https://www.lemonde.fr/planete/article/2020/04/24/nous-attendons-d-etre-contactes-par-l-ars-mais-il-ne-se-passe-rien-le-fiasco-des-tests-en-fra6037647_3244.html.

Sherry Glied, Peter C. Smith, David M. Cutler, Adriana Lleras-Muney, and Tom Vogl. Socioeconomic status and health: Dimensions and mechanisms, 09 2012.

Hervé Le Bras. L’é pidé mie et son terrain social, June 2020. URL https://jean-jaures.org/nos-productions/l-epidemie-et-son-terrain-social.LibraryCatalog:jean-jaures.org.

Piotr Lewandowski. Occupational exposure to contagion and the spread of covid-19 in europe. 2020.

John McLaren. Racial disparity in covid-19 deaths: Seeking economic roots with census data. Technical report, National Bureau of Economic Research, 2020.

Laura Montenovo, Xuan Jiang, Felipe Lozano Rojas, Ian M Schmutte, Kosali I Simon, Bruce A Weinberg, and Coady Wing. Determinants of disparities in covid-19 job losses. Technical report, National Bureau of Economic Research, 2020.

Ronald Oaxaca. Male-Female Wage Differentials in Urban Labor Markets. International Economic Review, 14(3):693–709, 1973. ISSN 0020-6598. doi: 10.2307/2525981. URL https://www.jstor.org/stable/2525981. Publisher: [Economics Department of the University of Pennsylvania, Wiley, Institute of Social and Economic Research, Osaka University].

Nicholas W Papageorge, Matthew V Zahn, Michèle Belot, Eline van den Broek-Altenburg, Syngjoo Choi,Julian C Jamison, Egon Tripodi, et al. Socio-demographic factors associated with self-protecting behavior during the covid-19 pandemic. Technical report, Institute of Labor Economics (IZA), 2020.

Claudia Persico and Kathryn R Johnson. The effects of increased pollution on covid-19 cases and deaths. Available at SSRN 3633446, 2020.

Matthew A. Raifman and Julia R. Raifman. Disparities in the Population at Risk of Severe Illness From COVID-19 by Race/Ethnicity and Income. American Journal of Preventive Medicine, 59(1):137–139, July 2020. ISSN 0749-3797. doi: 10.1016/j.amepre.2020.04.003. URL https://www.ncbi.nlm.nih.gov/pmc/articles/PMC7183932/.

Max Roser, Hannah Ritchie, Esteban Ortiz-Ospina, and Joe Hasell. Coronavirus disease (covid-19)– statistics and research. Our World in data, 2020.

Filipa Sa. Socioeconomic determinants of covid-19 infections and mortality: Evidence from england and wales. Technical report, Institute of Labor Economics (IZA), 2020.

Stefano Scarpetta, Mark Pearson, Francesca Colombo, and Frederico Guanais. Testing for covid-19: A way to lift confinement restrictions. OECD Policy Responses to Coronavirus (COVID-19), May 2020. URL https://www.oecd.org/coronavirus/policy-responses/testing-for-covid-19-a-way-to-lift-confinement-restrictions-89756248/#boxsection-d1e31.

Justin D Silverman, Nathaniel Hupert, and Alex D Washburne. Using influenza surveillance networks to estimate state-specific prevalence of sars-cov-2 in the united states. Science translational medicine, 2020.

SPF. Évaluation de la surmortalité pendant les canicules des é té s 2006 et 2015 en france mé tropolitaine, April 2019. URL https://www.santepubliquefrance.fr/content/download/209360/2415511.

Emily E Wiemers, Scott Abrahams, Marwa AlFakhri, V Joseph Hotz, Robert F Schoeni, and Judith A Seltzer. Disparities in vulnerability to severe complications from covid-19 in the united states. Technical report, National Bureau of Economic Research, 2020.

Zunyou Wu and Jennifer M McGoogan. Characteristics of and important lessons from the coronavirus disease 2019 (covid-19) outbreak in china: summary of a report of 72 314 cases from the chinese center for disease control and prevention. Jama, 323(13):1239–1242, 2020.

Vasil I Yasenov. Who can work from home? 2020.

